# Controlling risk of SARS-CoV-2 infection in essential workers of enclosed food manufacturing facilities

**DOI:** 10.1101/2021.05.14.21257244

**Authors:** Julia S. Sobolik, Elizabeth T. Sajewski, Lee-Ann Jaykus, D. Kane Cooper, Ben A. Lopman, Alicia NM. Kraay, P. Barry Ryan, Juan S. Leon

## Abstract

The SARS-CoV-2 global pandemic poses significant health risks to workers who are essential to maintaining the food supply chain. Using a quantitative risk assessment model, this study characterized the impact of risk reduction strategies for controlling SARS-CoV-2 transmission (droplet, aerosol, fomite-mediated) among front-line workers in a representative enclosed food manufacturing facility. We simulated: 1) individual and cumulative SARS-CoV-2 infection risks from close contact (droplet and aerosols at 1-3m), aerosol, and fomite-mediated exposures to a susceptible worker following exposure to an infected worker during an 8h-shift; and 2) the relative reduction in SARS-CoV-2 infection risk attributed to infection control interventions (physical distancing, mask use, ventilation, surface disinfection, hand hygiene). Without mitigation measures, the SARS-CoV-2 infection risk was largest for close contact (droplet and aerosol) at 1m (0.96, 95%CI: 0.67–1.0). In comparison, risk associated with fomite (0.26, 95%CI: 0.10–0.56) or aerosol exposure alone (0.05, 95%CI: 0.01–0.13) at 1m distance was substantially lower (73-95%). At 1m, droplet transmission predominated over aerosol and fomite-mediated transmission, however, this changed by 3m, with aerosols comprising the majority of the exposure dose. Increasing physical distancing reduced risk by 84% (1 to 2m) and 91% (1 to 3m). Universal mask use reduced infection risk by 52-88%, depending on mask type. Increasing ventilation (from 0.1 to 2-8 air changes/hour) resulted in risk reductions of 14-54% (1m) and 55-85% (2m). Combining these strategies, together with handwashing and surface disinfection, resulted in <1% infection risk. Current industry SARS-CoV-2 risk reduction strategies, particularly when bundled, provide significant protection to essential food workers.

**Significance Statement:** Using mathematical modeling, we find that workers in enclosed food manufacturing facilities are at higher risk of SARS-CoV-2 infection from close contact transmission (exposure to large droplets and small aerosol particles) than fomite transmission. Thus, strategies protecting workers should prioritize close contact transmission pathways, such as physical distancing, universal mask use, and room air changes, with surface disinfection (reducing fomite transmission) and handwashing of secondary importance. Our work supports current international (EU-OSHA), domestic (FDA, OSHA), and food industry-standard guidance for managing COVID-19 transmission in essential workers in the food manufacturing sector. Although our model was designed for an indoor food manufacturing setting, it can be readily adapted to other indoor environments and infectious respiratory pathogens.

## Introduction

Essential food worker populations have been disproportionately affected by severe acute respiratory syndrome coronavirus 2 (SARS-CoV-2) illness and death (1, 2). In a survey of essential workers in California, the highest excess mortality increase (39%) was among food and agriculture workers (3). Distinctive food production and processing occupational hazards may increase SARS-CoV-2 transmission, including inadequate physical distancing (<2m), shared workspaces, and extended exposure durations (8-12 hour shifts) (2, 4). Protecting the health and safety of food workers is paramount for maintaining global food supply chains and consumer food security (5).

In the food industry, the relative importance of SARS-CoV-2 transmission pathways have not been quantified. Exposure to SARS-CoV-2 within food manufacturing (processing) facilities may occur through direct (droplet and aerosol) and indirect (fomite-mediated) transmission pathways (6). Droplet transmission is defined as close contact (<2m) exposure to large, virus-containing particles (>100um diameter) generated by open-mouth respiratory events (e.g. coughing or sneezing) (7) that rapidly fall to the floor and/or nearby surfaces. Nearby susceptible individuals may be infected by SARS-CoV-2 through direct infectious droplet spray onto their mucous membranes (eyes, nose, mouth) or inhalation into the upper airways. Several large outbreaks in meat and poultry facilities (1, 2, 8) have occurred in which workers were in close contact (<2m) for extended durations, suggesting droplet transmission may be a key driver of SARS-CoV-2 infection risk in these settings.

In contrast to large droplets, aerosol transmission is associated with inhalation of small particles into the upper and lower respiratory tract. Small aerosol particles are historically defined as <5-10 µm in diameter, but recently recognized to include a wider range of particle sizes (<100 µm) (9, 10). Given the continuous range of particle sizes (11), the differentiation between aerosol and droplet sizes as defined by specific cut-off points likely remains debatable. Exposure to these aerosol particles can occur both in close contact and at further distances (up to 9m) (11). Aerosol particles are secreted during all respiratory events, especially during breathing and speaking (12, 13), and epidemiologic studies suggest viral accumulation and persistence during large indoor gatherings with poor ventilation (14-19). For example, SARS-CoV-2 RNA was detected in air samples from hospitals (20, 21) and in empirical laboratory studies, demonstrating that viable virus can remain suspended in the air for several hours (22, 23). Moreover, high viral shedding, including prior to symptom onset (24, 25), has important implications for both droplet and aerosol transmission.

Compared to direct transmission, indirect transmission via contaminated fomite surfaces (26-28) is considered less common, but possible, for SARS-CoV-2 (29, 30). For example, SARS-CoV-2 viral RNA has been detected on surfaces in various settings (21, 31-33) and laboratory studies found variable persistence of virus on fomites (up to 72h) across different surface types (22). While detection of RNA is not indicative of infectious virus (34) and laboratory conditions may not reflect viral persistence in real life scenarios (35, 36), frequent food worker tactile events and shared workstations warrant investigating the role of fomite-mediated transmission in food production and processing settings.

U.S. and international regulatory bodies (U.S. Occupational Safety and Health Administration [OSHA], European Union-OSHA), food safety agencies (USDA, FDA) and the food industry have issued infection control guidance for worker safety in food production and processing facilities, including symptom screening, physical distancing, mask use, and enhanced surface disinfection and handwashing practices (37-39). These measures pair with existing food safety measures under the FDA’s Food Safety Modernization Act, which provide standards for sick worker furlough, surface disinfection, and hand hygiene (handwashing, glove use) (40). Empirical and modeling studies suggest physical distancing (41, 42), mask use (43-47), and hand washing and surface disinfection (48-50) are effective measures against SARS-CoV-2 transmission. However, evidence is lacking to guide the food industry on the relative importance of these interventions on SARS-CoV-2 transmission among essential workers within food manufacturing facilities.

Quantitative microbial risk assessment (QMRA) is a mathematical modeling framework used to evaluate health risks associated with direct and indirect transmission pathways and to provide insight into efficacy of infection control strategies. Commonly applied in the food and environmental safety sectors (51-55), QMRA models have recently been used to characterize SARS-CoV-2 risk in healthcare (18, 56, 57), wastewater treatment facilities (58, 59), and community-based fomite transmission (50, 60, 61), but not yet in the food manufacturing setting. In this study, a stochastic QMRA model was used to quantify the impact of risk reduction measures (physical distancing, masking, ventilation, surface disinfection, and hand hygiene) for controlling SARS-CoV-2 transmission (droplet, aerosol, fomite-mediated) among essential (front-line) workers in a representative enclosed food manufacturing facility. This work advances the evidence-base for effective risk mitigation strategies currently implemented by the food industry and can be used to inform best practices for protecting essential workers.

## Results

### 3.1 Relative contribution of SARS-CoV-2 transmission routes in enclosed food manufacturing facilities with a coughing infected worker

Assuming a symptomatic infected worker (cough frequency ranged from 10-39 coughs per hour [63, 109]), we investigated the relative contribution of each transmission route (aerosol, droplet, fomite) by the distance travelled for each size class of expelled infectious particles for 1h and cumulative 8h exposures (Table 1). At 8h exposure and 1m distance between the infected and susceptible individuals, droplets (≥ 50µm) contributed 90% (absolute number: 478, 95% CI: [156-1,460]) of the infectious viral load, followed by aerosols (< 50µm) contributing 1.3% (absolute number: 7.0, 95% CI: [2-21]) and the remaining 8.3% of the infectious virus load coming from droplet and aerosol fall-out onto fomites (absolute number: 44, 95% CI: [16-122]). At 2 and 3m, the relative contribution of each transmission route to infectious virus load shifted to favor aerosols (31-59%) and fomites (25-48%), although the fomite and aerosol absolute viral load at 2 and 3m (3-11 PFUs) was 91-99% lower than the droplet absolute viral load at 1m (478 PFU). The patterns for 1h were similar (Table 1). Thus, the relative contribution of each mechanism of infectious particle spread was influenced by distance, with infectious droplet transmission representing the largest contribution to dose at 1m.

**Table 1.** Relative contributions (as a percentage) of droplet, aerosol, and fomite-mediated transmission modes to the cumulative SARS-CoV-2 viral load as a function of distance from initiating transmission events (coughing) and exposure time (1h or 8h).

| Time (h) | 1m | 2m | 3m | Combined viral load (PFU) ≤3m |
| --- | --- | --- | --- | --- |
| **1h** |  |  |  |  |
| Droplet | 98.4% | 47.9% | 31.2% | 60.6 |
| Aerosol | 0.8% | 42.4% | 64.7% | 1.5 |
| Fomite | 0.8% | 9.7% | 4.1% | 0.6 |
| **Absolute viral load (total PFU) at each distance** | 60.7 | 1.2 | 0.8 |  |
| **8h** |  |  |  |  |
| Droplet | 90.4% | 20.6% | 16.4% | 484.5 |
| Aerosol | 1.3% | 31.4% | 58.6% | 21.0 |
| Fomite | 8.3% | 47.9% | 25.0% | 57.5 |
| **Absolute viral load (total PFU) at each distance** | 528.8 | 22.2 | 11.9 |  |

Combining aerosol, droplet, and fomite-mediated transmission pathways resulted in combined risk estimates for exposures at near distances (1m, 2m, and 3m) and beyond (>3m, aerosols only, Figure 1B). Considering combined risks associated with close contact transmission (≤3m) at cumulative 8h exposures, 1m distance (0.98, 95% CI: [0.76-1.0]) resulted in the greatest risk, followed by 2m (0.15, 95% CI: [0.07-0.32]) and 3m (0.09, 95% CI: [0.04-0.18]) distances. Infection risks associated with aerosol and droplet transmission, without fomite-mediated transmission, remained elevated at 1m: (0.96, 95% CI: 0.67–1.0). Aerosol transmission alone resulted in substantially lower combined infection risks (8h: 0.05, 95% CI: [0.01-0.13]) (Figure 1B). The combined infection risk associated with small, aerosolized particles (<50µm) resulting from closed mouth, nasal breathing events (8h: 2×10^−4^, 95% CI: [6×10^−5^ – 6×10^−4^]) was smaller than the risk from aerosolized particles (<50µm) resulting from coughing events (8h: 0.05, 95% CI: [0.014-0.13]) (*SI Appendix*, Figure S2). While we initially intended to conduct all of our analyses with either a symptomatic (coughing) or asymptomatic (breathing) infected worker, given the nominal combined infection risk for breathing in the absence of any interventions, we determined coughing events appear to drive the risk within the context of this simulated manufacturing facility. Thus, we proceeded with only a symptomatic (coughing) infected worker for analyses moving forward. Fomite-mediated transmission associated with direct tactile events with a work surface contaminated from aerosol and droplet virus fallout resulted in modest infection risks (8h): ranging from 1m (0.26, 95% CI: [0.10-0.56]), 2m: (0.07, 95% CI: [0.02-0.20]), 3m (0.02, 95% CI: [0.006-0.06]), and >3m (7.2 × 10^−11^, 95% CI: [1.4 × 10^−11^ – 1.1 × 10^−10^]). As anticipated, combined risks as well as risks associated with aerosol and fomite-mediated transmission accumulated with increasing exposure time from 1 to 8 hours (Figure 1B). In particular, combined risks at 1m distancing started to plateau near 5h exposure and appeared to reach the exponential dose-response upper bound of the probability of infection equal to 1.0 after 8h of cumulative exposure from a coughing infected worker. Considerable variability in the infection risk estimates were noted, as represented by wide 95% confidence intervals (*SI Appendix*, Table S1). This is consistent with our sensitivity analyses (*SI Appendix*, Table S3), which identified variability propagating through the model associated with only a few parameters (i.e. virus titer in saliva, cough frequency, inhalation and deposition rate), all of which were found to be strongly positively correlated with infection risk.

**Figure 1.**
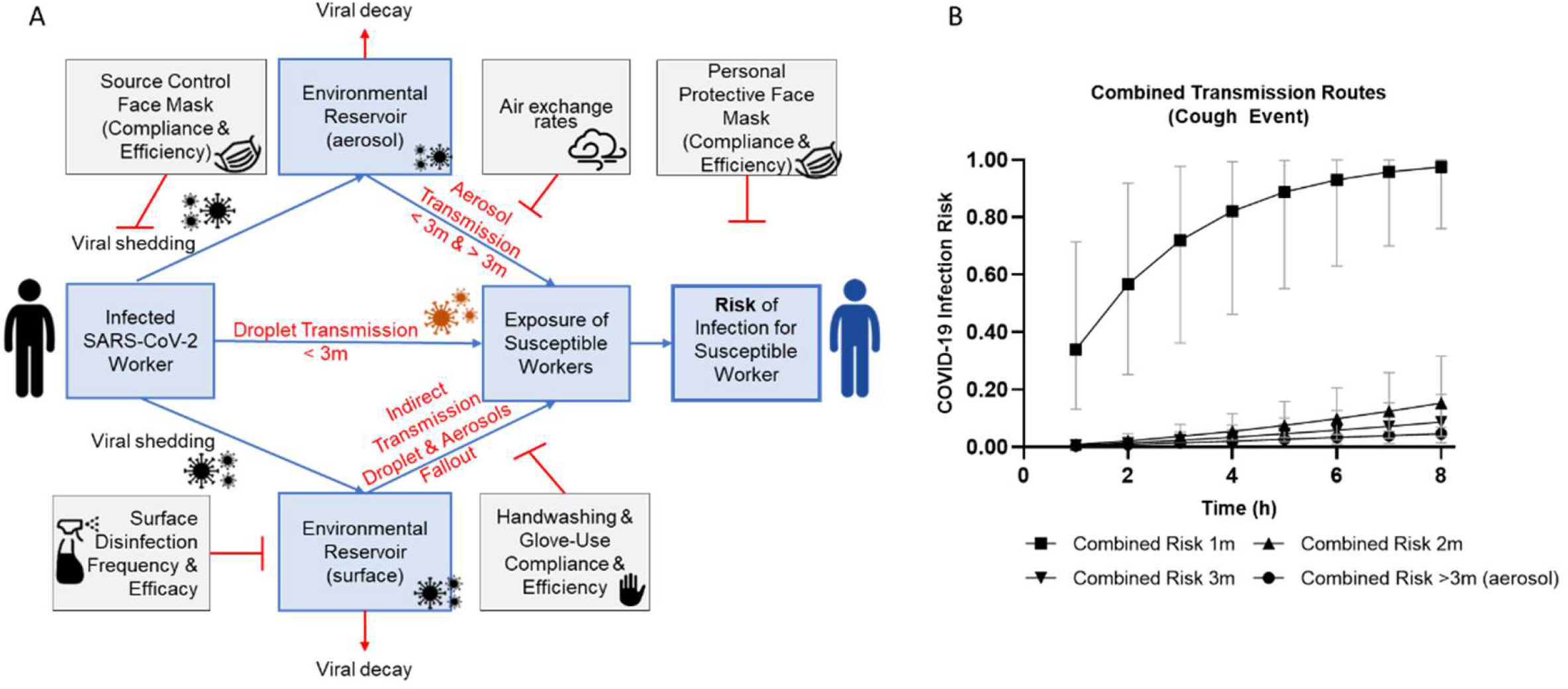
A-B. SARS-CoV-2 QMRA schematic for respiratory event (coughing versus breathing) and infection risk through aerosol, close contact (1-3m, droplet and aerosol), and fomite-mediated transmission assuming no risk mitigation interventions. A. This conceptual model depicts the three transmission pathways (close contact [droplet and aerosol], aerosol, and fomite-mediated) within a representative food manufacturing facility, initiating with a single infected worker either coughing (symptomatic) or breathing (asymptomatic) to generate virus-containing respiratory droplets and aerosols. Droplets fall rapidly due to gravitational forces and were categorized by size and distance traveled from source based on empirical experiments and modeling studies (75, 110): <1m (50-750 μm), 1-2m (50-100 μm), and 2-3m (50-60 μm). Aerosols were defined as <50 μm in diameter with the ability to become aerosolized and remain suspended in the air throughout the entire facility space. B. Infection risk from combined transmission events (aerosol, droplet, fomite-mediated) in association with exposure to an infected worker (coughing) over a period of 1-8 h and as a function of distance.

### 3.2 Comparison of R_0_ values derived from SARS-CoV-2 combined transmission risks

To assess the order of magnitude of these SARS-CoV-2 infection risk estimates at the population-level, using a 7-day infectious period, we converted the combined transmission risk estimates to population-level R_0_ values. At 1m, R_0_ values ranged from 2.38 (1h) to 6.83 (8h) (*SI Appendix*, Table S4). As all combined risk estimates (exposures ranging from 1-8h) at 1m distancing, when converted to R_0_ values, were >1, this suggests that SARS-CoV-2 transmission would be sustained when workers are in close contact to each other. An R_0_ value of 1.07 (95% CI: 0.48-2.2) was estimated for 2m exposures with 8h cumulative risks. Distances of 3m and beyond were found to have all R_0_ values <1. These model derived R_0_ values were consistent with reported population-level disease transmission events with R_0_ values ranging from: 2.3 to 11 (62-64). Despite heterogeneity across attack rates and outbreak scenarios, we found that our risk estimates, when scaled, were representative of observed SARS-CoV-2 transmission dynamics at the population level.

### 3.3 Impact of individual interventions targeting combined risk

Increasing physical distancing resulted in substantial combined risk reductions following 1h exposure. An additional 1m distancing from 1 to 2m led to a 97% reduction in risk and from 2 to 3m led to a 35% reduction in risk. Increasing physical distancing by 2m from 1 to 3m resulted in a 98% risk reduction (Figure 1B, *SI Appendix*, Table S2). A similar trend in risk reductions was found following an 8h exposure: an additional 1m distancing from 1 to 2m led to a 84% reduction in risk and from 2 to 3m led to a 43% reduction in risk. Increasing physical distancing by 2m from 1 to 3m resulted in a 91% risk reduction. These analyses demonstrated that physical distancing 2m and beyond provided the greatest relative reduction in combined infection risk.

Universal mask use at 1m distance following 8h cumulative exposure with a coughing infected worker reduced combined infection risk by 52% (cloth mask: 0.47 risk, 95%CI: 0.20–0.87), 64% (surgical mask: 0.35 risk, 95%CI: 0.12–0.76), 88% (double masking [surgical followed by cloth]: 0.12 risk, 95%CI: 0.03– 0.55), and 99% (N95 respirator: 0.01 risk, 95%CI: 0.004–0.02), relative to no mask use (Table 2), leading to absolute risk of infection at 1m of 0.01-0.47, depending on mask type. Combining mask use and physical distancing resulted in enhanced risk reduction, suggesting a synergistic effect between these two interventions. For example, following 8h cumulative exposure, distancing from 1-2m paired with mask use resulted in risk reductions of 91% across all mask types. Physical distancing from 2-3m resulted in risk reductions ranging from 53-61% reduction depending on the mask type. Physical distancing from 1-3m resulted in risk reductions ranging from 96-97% reduction depending on the mask type. In all cases with the exception of N95 respirators, physical distancing paired with mask use resulted in a greater risk reduction than distance or mask use alone.

**Table 2.** Impact of mask use on combined infection risk following 8h cumulative exposure to an infected worker (coughing) as a function of distance.

**Impact of mask interventions on infection risk (95% CI) and percent risk reduction (%)**
|  |  | 1m | 2m | 3m | Aerosols | Average<br>Percent<br>Risk<br>Reduction |
| --- | --- | --- | --- | --- | --- | --- |
| **No Mask** | Risk (95% CI) | 0.98 (0.76-0.99) | 0.15 (0.07-0.32) | 0.09 (0.04-0.18) | 0.05 (0.01-0.13) |  |
|  | --- | --- | --- | --- | --- | --- |
| **Cloth** | Risk (95% CI) | 0.47 (0.20-0.87) | 0.04 (0.02-0.10) | 0.02 (0.007-0.05) | 0.007 (0.001-0.03) |  |
|  | % Reduction | 51.8% | 73.8% | 79.0% | 85.3% | 72.6% |
| **Surgical** | Risk (95% CI) | 0.35 (0.12-0.76) | 0.03 (0.01-0.09) | 0.01 (0.005-0.04) | 0.004 (0.0004-0.02) |  |
|  | % Reduction | 64.1% | 80.4% | 83.6% | 90.5% | 79.1% |
| **Double mask\*** | Risk (95% CI) | 0.12 (0.03-0.55) | 0.01 (0.002-0.054) | 0.004 (0.001-0.021) | 0.001 (0.000-0.01) |  |
|  | % Reduction | 87.7% | 93.3% | 95.4% | 97.7% | 93.7% |
| **N95** | Risk (95% CI) | 0.01 (0.004-0.02) | 0.0009 (0.0003-0.002) | 0.0004 (0.0001-0.0009) | 0.0001(0.00003-0.0003) |  |
|  | % Reduction | 98.9% | 99.4% | 99.6% | 99.8% | 99.4% |

\*Double masking defined as surgical mask layered underneath a cloth mask.

To evaluate the impact of ventilation on combined infection risk following an 8h exposure with an infected worker, we increased air change rates per hour from 2 to 8 ACH (Figure 2). Compared to baseline (ACH 0.1), increasing air change to between 2 to 8 ACH resulted in a percent reduction in combined infection risk at: 1m (mean of 2-8 ACH: 36%, range: 14%-54%), 2m (mean of 2-8 ACH: 74%, range: 55%-85%), 3m (mean of 2-8 ACH: 77%, range: 60%-87%), and >3m (mean of 2-8 ACH: 82%, range: 69%-90%). Similar to mask usage, reductions in the combined infection risk were enhanced when the susceptible worker was 2m or 3m, compared to 1m, distance away from the infected worker in the presence of increasing air exchange. Following 8h cumulative exposure, distancing from 1-2m resulted in risk reductions of 92-95% for ACH ranging from 2-8. Physical distancing from 2-3m produced risk reductions ranging from 50-51% for 2-8 ACH. Combining the two physical distancing ranges (1-3m) produced risk reductions ranging from 96-98% for 2-8 ACH. Rank prioritizing these single interventions suggests physical distancing, followed by mask use, and then increasing facility ventilation results in the largest combined risk reductions to a susceptible worker after an 8h-shift with a coughing infected worker. However, the impact of distancing from 2m and beyond was notably enhanced when paired with mask use (any mask type) or air change rates (≥ 2 ACH), which resulted in combined risk reductions >91%.

**Figure 2.**
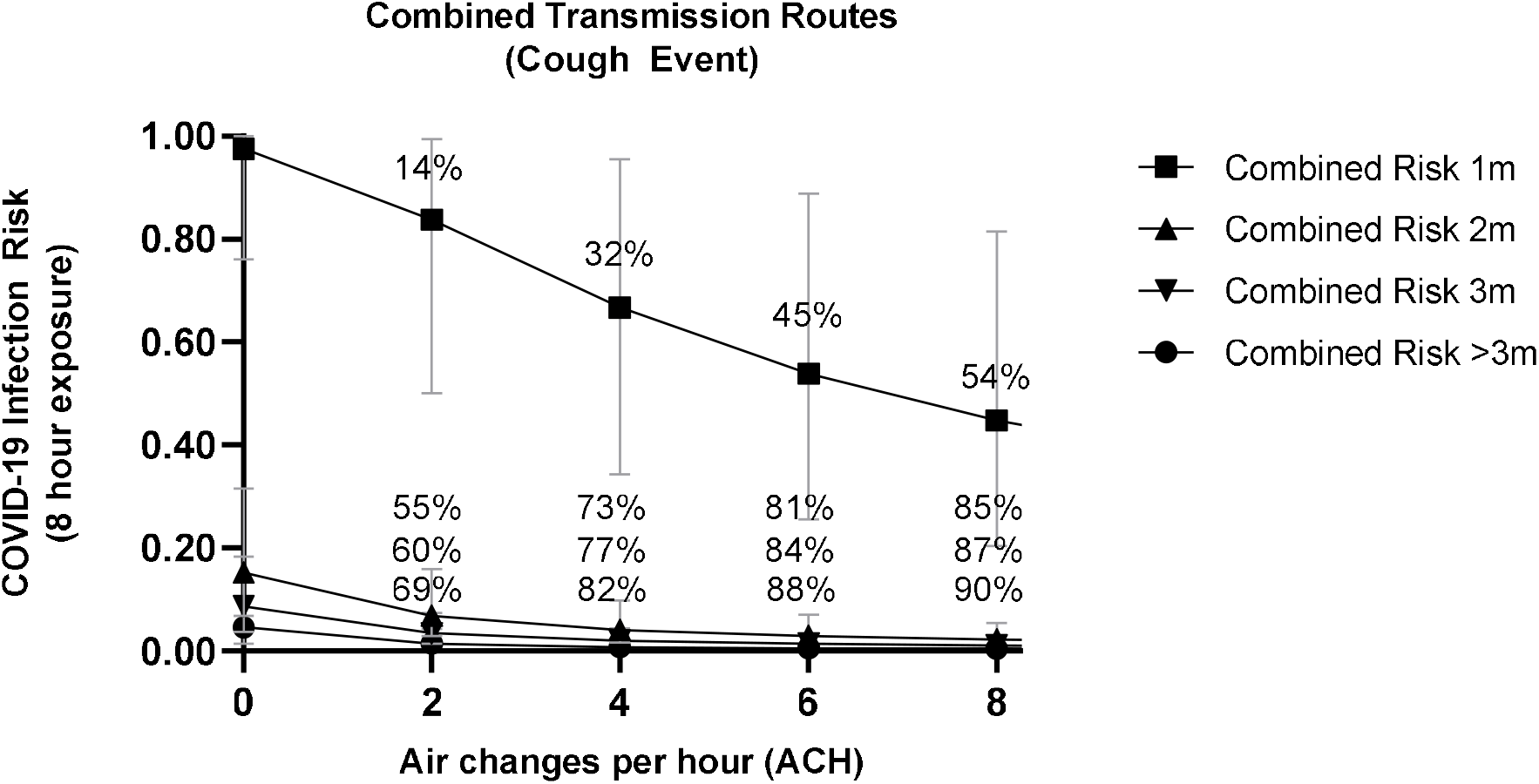
Impact of increasing air exchange on combined infection risk reduction following an 8h-exposure to an infected worker (coughing) at various distances. For reference, air changes per hour (ACH) of 2-6 are representative of typical indoor food manufacturing facilities based on survey results. Included percentages represent the percent reduction in SARS-CoV-2 infection risk relative to no air exchange (baseline ACH = 0.1) for combined risk at the four distances (1m, 2m, 3m, and >3m) modeled.

### 3.4 Impact of individual interventions targeting fomite-mediated risk

Although the relative contribution of fomites to infectious dose was low compared to droplet transmission (Table 1), fomite-mediated transmission does contribute to worker risk, particularly at close exposures. Thus, we investigated interventions specifically targeting fomite-mediated, 8h cumulative exposures from a coughing infected worker at 1m distance (worst-case scenario). Similar to the close contact and aerosol transmission pathways, mask use effectively reduced fomite-mediated transmission. Following 8h cumulative exposure at 1m, mask use reduced fomite-mediated infection risk by 62% (cloth mask: 0.10 risk, 95% CI: [0.03–0.25]), 63% (surgical mask: 0.09 risk, 95% CI: [0.03–0.25]), 88% (double masking [surgical followed by cloth]: 0.03 risk, 95% CI: [0.005–0.15]), and 99% (N95 respirator: 0.003 risk, 95% CI: [0.001–0.008]), relative to no mask use. Handwashing (2 log_10_ virus removal efficacy) and hand sanitizer (3 log_10_ virus removal efficacy) use resulted in large fomite-mediated risk reductions relative to no intervention: handwashing hourly (98.8% reduction; 0.003 risk, 95% CI: [0.001–0.008]) and alcohol-based hand sanitizer hourly (99.88% reduction; 3×10^−4^ risk, 95% CI: [1×10^−4^ –8×10^−4^]) (*SI Appendix*, Figure S3). Hourly glove changes (assuming handwashing completely removed all viral contamination and was performed before donning clean gloves) mitigated all SARS-CoV-2 fomite-mediated risk (data not shown). Only hourly surface disinfection (the most frequent surface disinfection scenario tested) resulted in fomite-mediated risk reductions comparable to those achieved by handwashing: hourly surface disinfection (3log_10_ virus removal efficacy) (99.96% reduction; 1×10^−4^ risk, 95% CI: [3×10^−5^ –3×10^−4^]) and hourly surface disinfection (4log_10_ virus removal) (99.99% reduction; 1×10^−5^ risk, 95% CI: [3×10^−6^ –3×10^−5^]), relative to no intervention (*SI Appendix*, Figure S3).

### 3.5 Impact of bundled risk mitigation strategies targeting combined infection risk

Our survey of food manufacturing facilities and discussions with food industry experts (data not shown) indicated that various, simultaneous COVID-19 specific infection control measures (mask use, physical distancing, air changes per hour, hand hygiene, surface disinfection, sick worker furlough) have been consistently implemented in food production and processing facilities, in practice. Bundled strategies of physical distancing of 2m, universal mask wearing (cloth, surgical, or double masking), and at least 2 ACH (industry standard), combined with handwashing (hourly) and surface disinfection (twice per shift, 4h and 8h) resulted in combined infection risks less than 1% in an 8h-shift (Table 3). Comparable combined infection risks (<1%) were achieved with bundled strategies incorporating reduced frequencies of surface disinfection (once per shift) and hand hygiene (twice per shift); reduced efficacies of surface disinfection (1 to 3 log_10_ virus removal efficacy); and with hand sanitizer used interchangeably with handwashing (data not shown), in addition to mask use and ACH. If physical distancing is not possible, at 1m distancing, the largest combined infection risk reduction of 98% resulted from double mask use with 6 ACH (risk: 0.02, 95% CI: [0.002-0.14]), relative to no interventions.

**Table 3.**
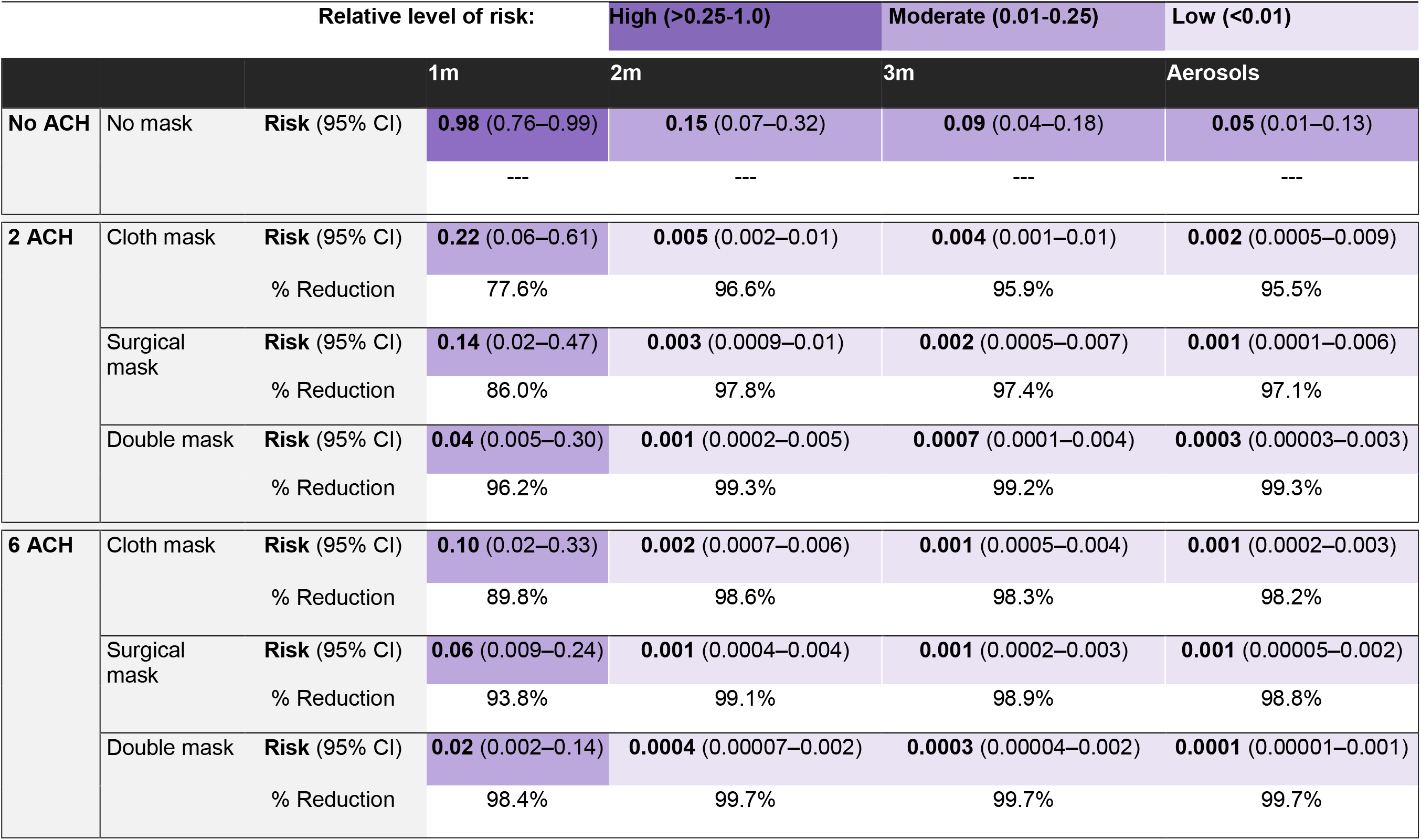
Impact of bundled interventions (mask use, ventilation, hourly handwashing and surface disinfection twice per shift [4h and 8h]) on infection risk (95% CI) and percent risk reduction (%) following 8h cumulative exposure to an infected worker (cough event as a function of distance). Colors indicate risk level from each bundled package. Dark purple indicates a high relative level of risk (>0.25 – 1.0), medium purple indicates a moderate relative level of risk (0.01-0.25), and light purple indicates a low relative level of risk (<0.01).

### 3.6 Sensitivity analyses

Sensitivity analyses were conducted to identify the most influential parameters for SARS-CoV-2 infection risk, reported as Spearman’s correlation coefficients. The parameters with the greatest contribution to infection risk were consistent across the aerosol and droplet transmission pathways and included the SARS-CoV-2 titer in saliva (aerosol ρ=0.58; droplet ρ= 0.73); cough frequency of the infected worker (aerosol ρ=0.24; droplet ρ=0.30); inhalation rate (aerosol ρ=0.19; droplet ρ= 0.16); and lung deposition rate (aerosol ρ=0.21; droplet ρ=0.16) (*SI Appendix*, Figure S1). The dose-response *k* parameter was found to have a moderate correlation with SARS-CoV-2 infection risk (droplet ρ = 0.36). The parameters with the greatest impact on reducing infection risk included mask filtration efficiency (ρ= -0.13 to -0.16)) and the number of air changes per hour (ρ= -0.52 to -0.35). Notably the parameters describing tactile events, such as transfer efficiency and number of contacts, generally had smaller effects (ρ = -0.002 to 0.005) on the infection risk estimates.

We also evaluated the contribution of individual parameters to the overall variability associated with the estimated infection risk. Variability, representing temporal, geographical, and individual heterogeneity, was generally quite low for each individual parameter, with the largest values associated with: (1) the distribution of particle sizes found in a cough or breathing event; (2) cough frequency; (3) masking efficacies; (4) room air exchange; (5) transfer efficiencies from fomite-to-hand; and (5) transfer efficiencies from hand-to-face mucous membranes. (*SI Appendix*, Table S3). The overall impact of variability on aerosol and droplet infection risk estimates, defined as the variability ratio (97.5^th^ model estimate/the median model estimate), were modest (variability ratio: 6.8 to 10.2). For the fomite-mediated transmission pathway, considerable variability was found to propagate through the simulated viral concentration on the fomite surface, to the viral concentration on hands, and ultimately for the fomite-mediated risk.

## Discussion

In this study, a stochastic quantitative risk assessment model was used to quantify the impact of risk reduction strategies (physical distancing, masking, ventilation, surface disinfection, and hand hygiene) for controlling SARS-CoV-2 transmission (droplet, aerosol, fomite-mediated) among essential (front-line) workers in a representative enclosed food manufacturing facility. Collectively, our modeling results indicate that droplets were the dominant transmission mode (90%) at 1m distance, delivering the highest infectious viral load to essential food workers during 8h-shifts, relative to aerosol and fomite-mediated routes (1-8%). In comparison, at distances 2m and beyond, the viral load shifted to aerosol and fomite-mediated transmission. However, their absolute dose contribution (aerosol viral load: 7.0, 95% CI: [2-21]; fomite-mediated viral load: 44, 95% CI: [16-122]) remained 91-99% lower than droplet-mediated transmission at 1m, and absolute risk from all pathways combined was far lower than risk at 1m. Among the individual interventions, physical distancing (2m and beyond) and mask use resulted in the largest risk reductions to a food worker. Bundled interventions (at least 2 ACH, 2m physical distancing, universal mask use, hand hygiene and surface disinfection) reduced essential food worker risk to below 1% for an 8h cumulative exposure in an enclosed food manufacturing facility.

The droplet-mediated transmission pathway at 1m contributed at least a 11-fold higher viral load than fomite-mediated or aerosol pathways at 1m (1h and 8h cumulative exposures), translating to a higher attributable risk of infection. Two mechanisms explaining this result include the particle size distribution generated during cough events and the delivery of the highest infectious dose in these cough-generated particles. While cough respiratory events generate both small and large particles, large particles (droplets >50 µm) represent a higher proportion of the particles released while coughing relative to other respiratory events (65). These large droplets, which have a larger volume, comprise the majority (99.9%) of the fluid volume expelled during a cough event (66). While empirical work has shown influenza virus-infected patients generate small aerosols (<5 μm) containing viable virus (67-70), to date, the distribution of infectious SARS-CoV-2 in aerosols and droplets has not been validated. Using a Poisson distribution to estimate the number of viruses enclosed in a droplet, Wang *et al*., estimated that upon particle emission, viruses were mostly contained in droplets (>10 μm). However, these particle sizes were predicted to reduce to approximately 2 μm in size following evaporation (71). While additional empirical studies are needed to confirm the size-dependent concentration and infectivity of SARS-CoV-2 in respiratory particles, we chose to implement a conservative approach by assuming uniform virus concentration across particle sizes. We speculate this approach could potentially lead to heightened “worst-case scenario” risk estimates associated with close contact droplet exposures. However, the efficacy of interventions and the relative risk reductions associated with these interventions should remain reliable as any potential biases introduced due to virus concentration across particle sizes would be incorporated into both the intervention and control risk estimates (72).

Although our model showed that droplets (50-750 μm) played a larger role in close contact (1m) transmission, Azimi *et al*. reported no significant difference between larger respiratory droplets (>5 to 10 μm) (median: 40%, mean: 50%) and smaller respiratory aerosols (<10 μm) (median: 60%, mean: 50%) and their contribution to SARS-CoV-2 transmission aboard a cruise ship prior to passenger quarantine (p=0.32). Differences in modeling certain parameters, i.e., the proportion of aerosols to droplets emitted during a respiratory event and estimating the infectious dose, rather than using the existing SARS-CoV dose-response model, likely explain differences in findings. For our model, we determined that any uncertainty or variability introduced by extrapolating from a SARS-CoV dose-response to SARS-CoV-2 was nominal given the similarities between the viruses (high genetic and amino acid homology) (73) and shared transmission pathways. In a clinical setting, Jones *et al*., found that the relative contribution to overall infection risk was higher for aerosols 57% (33, 82%) versus droplet 35% (12, 55%) transmission (18). However, Jones *et al*. modeled a substantially smaller room volume (20-50 m^3^), incorporated continuous virus emission of aerosols (<10 μm) via exhaled breath in addition to aerosols and droplets generated from coughing, and assumed droplet particles have 50% the virus concentration of aerosols (compared to our assumption of equal virus concentration across particles). Such differences between the results of various risk modeling studies illustrate the sensitivity of risk estimates to key exposure parameters and the challenge in determining aerosol and droplet exposure attribution, which is likely context-specific.

While detection of SARS-CoV-2 on surfaces (74) continues to suggest the potential for fomite-mediated transmission, our model estimated only a very modest role for fomite-mediated transmission in enclosed food manufacturing facilities, with fomites accounting for <1% to 10% (1h exposure) of the viral load (Table 1). These results are in the range of those reported in other studies across diverse settings. In a clinical setting, fomite-mediated transmission was the lowest contributor to overall SARS-CoV-2 infection risk, representing 8.2% of the total transmission risk during a single patient care activity (18). Moreover, during a cruise ship outbreak, the median contribution of fomite transmission to infected cases was estimated to be 21% (17). Translating exposure dose to infection risk, our study also found fomite-mediated transmission resulted in low SARS-CoV-2 infection risks, as expected, following 1h exposure at 1m: 0.0033 95% CI: [0.001-0.009]. However, following 8h cumulative exposure at 1m, fomite-mediated risk increased to 0.26 95% CI: [0.10-0.56]. This infection risk translated to an R_0_ >1 (1.8, 95% CI: 0.71-3.9), which underscores the potential for increased risk with long exposure durations and suggests that in the situations for when droplet transmission risk is high, fomite transmission risk will also be high, potentially leading to competing or multiple pathway transmission dynamics. Our 1h fomite-mediated results are consistent with Wilson *et al*., who found fomite-mediated infection risk was approximately 1.0×10^−3^ for a single hand-to-fomite scenario with high SARS-CoV-2 bioburden and no surface disinfection (50). In agreement with the low contribution of fomite-mediated risk, Pitol *et al*., (60) and Harvey *et al*., (61) reported fomite-mediated risks associated with direct tactile events in community spaces (bus stations, gas stations, playgrounds) and on high touch non-porous surfaces (crosswalk buttons, trash can handles, door handles) ranging from 1.6×10^−4^ to 5.6×10^−9^. These lower risk estimates likely result from both studies simulating a single direct tactile event in a low prevalence community setting, whereas our model accounted for multiple tactile events over an 8h cumulative shift with a confirmed SARS-CoV-2 infected worker during their peak viral shedding period (20). Although heterogeneity in fomite-mediated infection risk is likely context-specific (i.e. depends on SARS-CoV-2 community prevalence, degree of surface contamination, types of initiating events [i.e., breathing vs. coughing], etc.), increasingly findings suggest that fomites are a relatively less important transmission pathway than are direct and indirect respiratory exposures.

The distance between infected and susceptible individuals was a major driver of infection risk in our model. Laboratory and modeling studies (75, 76) suggest that the highest concentrated exposures to both droplet and aerosol respiratory particles occur when within 1m of an infected individual. Consistent with these studies, our median SARS-CoV-2 infection risk was highest with close contact (droplet and aerosol) transmission at 1m with exposures ranging from 1h: (0.34 95% CI: [0.13-0.71]) to cumulative 8h: (0.96 95% CI: [0.67-0.99]). Similar findings have been reported in a clinical setting, with exposure in the “near-patient zone” resulting in a mean infection risk of 0.38 95%CI (0.18, 0.53) from a single patient care activity (18). Similarly, in a recreational setting, close contact transmission (droplet and aerosol exposures) was predominant over other transmission modes when individuals were restricted to their close quarter cruise ship cabins (∼14 sq. m) (17). Moreover, we found risk estimates decreased by over 80% when increasing the distance by 1m between the workers. This protective effect was consistent with previous studies finding that >1m physical distances were associated with large risk reductions and distances of >2m could be even more effective (42, 77).

In addition to distance, our findings suggest that facility space and layout also contribute to the relative and absolute risk of virus transmission to workers. For instance, at distances beyond large droplet exposures (>1m), we found a greater contribution of the infectious dose derived from the aerosol and fomite mediated transmission modes. These results align with reports of virus-laden aerosols (<50-100µm) capable of accumulating in the air over time in enclosed facilities (78). Despite this shift towards aerosol transmission at distances >1m, surprisingly, we found the relative infection risks associated with both aerosol and fomite-mediated transmission remained small. We hypothesize that large indoor spaces, like enclosed food manufacturing facilities, attenuate aerosol accumulation through dilution across the facility space. For example, a model of the seafood market in Wuhan found reduced median SARS-CoV-2 infection risks after 1 h exposure (2.23 × 10^−5^ [95% CI: 1.90 × 10^−6^ to 2.34 × 10^−4^]), likely due to aerosols disseminated over the >3000 cubic meter space (79). Similarly, our model represented a 1000 cubic meter facility, in which we assumed instantaneous mixing of the room air with homogenous distribution of aerosol particles throughout the facility. Re-running the model using a smaller space (37 cubic meters) produced a 17-fold increase in aerosol transmission-mediated risk (0.73, 95%CI: 0.33-0.98), suggesting small enclosed spaces (with minimal ventilation, ACH 0.1) may accelerate accumulation of virus-laden aerosols (data not shown). Taken together, these results highlight the importance of increased distance as an effective risk mitigation strategy and the need to contextualize facility spaces when discussing relative infection risks.

The risk reduction effect of universal mask use in our study is consistent with previous empirical (42, 77) and laboratory-based experiments (44-47). In our study, universal double masking (surgical mask layered underneath a cloth mask) by both the infected and susceptible workers reduced risk by 88 to 98% (1m to >3m). These findings are consistent with those reported by Brooks *et al*., where both the infected and susceptible individuals were fitted with double masks and the cumulative exposure to the susceptible individual was reduced by 96.4% (SD = 0.02) (80). Mask effectiveness is also dependent on individuals adhering to proper mask use (e.g. minimizing adjustment or touching the front of the mask, wearing the mask over the nose and mouth). Of course, proper mask use and adherence is necessary in order to realize the full benefit of masking in COVID-19 transmission risk reduction. While we assumed 100% mask compliance, this model could be used to evaluate varying levels of mask compliance and the impact on infection risk when only the infected worker uses a mask and not the susceptible worker, and vice versa.

Increasing ventilation in the facility, as expressed as the number of air changes per hour (ACH), resulted in a decreased infection risk, especially when combined with physical distancing and mask use. Consistent with recent work by Zhang et al. (79), Kennedy et al. (81), and Curtius *et al*. (82), these findings advance a growing body of evidence linking the association between ventilation, air movements in buildings, and the transmission of infectious diseases (measles, tuberculosis, influenza, and SARS) with primary respiratory exposure routes (83). Notably, poor room ventilation was specifically identified as a contributing factor in a SARS-CoV-2 outbreak in a German meat packing facility (84). A well-ventilated indoor room with frequent fresh air changes can prevent the accumulation of virus-laden aerosols (<50-100µm) (78), which have been demonstrated under controlled laboratory settings to remain suspended in the air for many seconds to hours (22); it also reduces the potential for super-spreader events (16, 85-87). Our findings advance the evidence-base of current recommendations to reduce aerosol-mediated transmission of SARS-CoV-2 (88, 89) which can be accomplished by increased ventilation (providing outdoor air to a space), avoiding air recirculation, and use of air cleaning and disinfection devices (88, 89). It is important to note, however, that the use of simpler interventions (masking and distancing) provided greater risk reduction than did increasing ACH alone, which would ultimately be a more expensive engineering investment. As most manufacturing facilities already implement 2-6 ACH as part of their standard operating procedures, prior to extensive ventilation investments, more study is needed on aerosol transmission dynamics and the value of targeted methods designed to increase ventilation in food manufacturing settings.

An important contribution of our work is quantitatively demonstrating that bundled interventions are highly effective infection control measures within enclosed food manufacturing facilities. Compliance with all standard recommendations (physical distance, universal mask usage, increased ventilation, handwashing, and surface disinfection) was found to reduce worker risk to under 1% in an 8h-shift. This work supports the efficacy of these bundled interventions. The high efficacy of the combined interventions involving masks and physical distancing is of particular interest given their relatively low cost, high-impact risk mitigation potential, and ease of scaling across diverse food manufacturing settings. Continued utilization of these two mitigation strategies is especially salient given uncertainties surrounding the emergence of SARS-CoV-2 variants, vaccine efficacy against the variants, and duration of immunity. Our model may be particularly helpful in informing decision-making for facility managers to prioritize which interventions to keep in place, and which to alter or stop, post-vaccination. The efficacy of advanced interventions, such as use of HEPA filtration (90) or far-UVC light inactivation (91) in controlling COVID-19 disease risk in this essential workforce can easily be incorporated in future simulations, as can the impact of various vaccination strategies.

Strengths of our model include a detailed exposure assessment design of food production and processing facilities; well-characterized human-environment interactions based on direct observation from prior fieldwork; vetting by industry and academic partners; and an extensive validation (e.g. scaling risk estimates to population-level R_0_) and calibration, steps that are not routinely done in QMRA models. This design enabled evaluating the relative contribution of each transmission pathway (droplet, aerosol, and fomite-mediated), generating combined risk estimates from SARS-CoV-2 exposures, and ultimately prioritizing evidence-based risk mitigation strategies informed by federal worker health and safety guidelines (92, 93) for food manufacturing facilities. A final strength of our model was incorporating new evidence from the field of aerosol physics to define aerosols and droplets beyond the classical cutoff of ∼10µm. Regarding limitations, the first is that at the time of this writing, there was no dose-response model specific to SARS-CoV-2. To address this, we leveraged the dose-response model for SARS and applied the upper 99.5% bound from 10,000 bootstrap iterations for the optimized *k* parameter. This translated into an ID_50_ dose of 100 infectious virus particles and falls within the documented ID_50_ range of 10-1000 infectious doses from animal and human studies for non-SARS-CoV and SARS (94). A second limitation was that few empirical studies have characterized respiratory particle distance traveled by size. Understanding the dynamics of infectious virus-laden respiratory particles at different distances, especially accounting for air transport dynamics within a respiratory event (11, 75), is paramount to characterizing risk and evaluating effective intervention strategies. Certainly, this is a rapidly evolving field and the model can be readily updated as new data emerge. A final limitation of this study is that virus contamination in the air, on fomite surfaces, and on hands was assumed to be homogenously distributed. Empirical studies on the distribution of respiratory particles in the air and on fomite surfaces following respiratory events are needed to further refine respiratory transmission pathway modeling.

Future research might include evaluation of SARS-CoV-2 risk associated with packaging in a food supply chain (95) and in outdoor agriculture production and harvest settings; and the impact of population-based interventions, including vaccination, testing, and worker furlough, on SARS-CoV-2 cases and mortality among this essential workforce. This model can also be adapted to other indoor settings (e.g. schools and daycares) to provide evidence-based guidance for SARS-CoV-2 risk mitigation strategies. While this model represented a food manufacturing facility encompassing frozen food processing and fresh cut produce packaging, given additional information on equipment and facility design, this model can readily be adapted to other prepared foods, meat, and poultry processing manufacturing facilities. Taken together, this work advances the evidence-base for existing global (39, 96), federal (37, 40) and food industry (97) guidelines as effective SARS-CoV-2 infection mitigation strategies to protect essential workers in food manufacturing facilities.

## Materials and Methods

### 2.1 Model overview

Model design of the indoor food manufacturing facility was informed by prior field studies conducted in fresh produce packing facilities along the southern United Sates and northern Mexican border states (98-101). As a representative food manufacturing facility, we also applied information on equipment and facility design from frozen food manufacturers associated with the frozen fruit and vegetable sector. In addition, we leveraged the modeling frameworks of SARS-CoV-2 aerosol transmission in a seafood market in Wuhan, China (79), Middle East Respiratory Virus (MERS) aerosol and droplet transmission in a hospital (102), and fomite-mediated transmission for influenza A virus (103, 104). Additional details on model vetting are provided in the *SI Appendix*.

### 2.2 Model structure

The overall model structure initiates with a single infected worker exposing a single susceptible worker over a work shift lasting up to 8h either through coughing (symptomatic) or breathing (asymptomatic) respiratory events. Virus-containing droplets and aerosols generated from these respiratory events then feed into the close contact, aerosol, and fomite-mediated transmission pathways. Please refer to *SI Appendix* for additional details on the transmission pathways by event. In our model, SARS-CoV-2 infection risk to the susceptible worker resulted from respiration/deposition of particles in the nasal-pharyngeal region (droplet and aerosol); direct spray onto mucous membranes (droplet); and indirect tactile transfers associated with contaminated fomite surface(s) (Figure 1). Exposure to aerosols (<50 µm) by the susceptible worker occurred both near (close contact 1-3m) and farther (beyond close contact >3m) from the infected individual. Exposure to droplets (50-750 μm) by the susceptible worker occurred only within close contact distancing. Here, droplet exposures were characterized by the transport properties of the droplets (i.e. ballistic gravitational trajectories) and the possible horizontal distance traveled. For instance, with a coughing infected worker, a susceptible worker at <1m distancing would be exposed to the full range of 50-750 μm droplets, whereas distancing of 1-2m would result in exposure to 50-100 μm droplets (droplets >100 μm having settled to the floor/fomite surface), and distancing of 2-3m would result in exposure to 50-60 μm droplets (droplets >60-100 μm having settled to the floor/fomite surface). In addition to these droplet exposures, when in close contact, a susceptible worker would also be exposed to aerosol particles (<50 µm), as coughing events produce both aerosol and droplet particles simultaneously. Viral contamination of a fomite surface occurred through respiratory particles falling from the air, either by the terminal settling velocity of aerosols or gravitational ballistic trajectory of droplets, onto a 0.5m by 0.5m stainless steel surface within 1m of the susceptible worker. Of the total respiratory particle fallout from the air, we assumed only a proportion of this total would land and contaminate the fomite surface (fomite surface area/cross-sectional facility area). Fomite-mediated transmission involved tactile contact between the susceptible worker’s fingers and palms (of both hands) and the fomite surface (accounting for surface area of the hand relative to the fomite surface); virus transfer from fomite to hands; followed by virus transfer from fingertips to facial mucous membranes (accounting for the surface area of the fingers relative to that of the hands). SARS-CoV-2 infection risk to the susceptible worker was calculated by incorporating the dose from each individual transmission pathway and applying an exponential dose-response model based on data from SARS-CoV and murine hepatitis virus infection in mice (105, 106). As there is currently no SARS-CoV-2 dose-response model, consistent with other SARS-CoV-2 QMRA models (18, 50, 60, 61), we applied this SARS-CoV dose-response given the high degree of comparability between SARS-CoV and SARS-CoV-2 (genetic and amino acid homology, transmission pathways, etc.) (73). Infection control measures were implemented to target one or more of the SARS-CoV-2 transmission pathways (droplet, aerosol, and fomite-mediated). For instance, mask use by the infected worker aimed to interrupt the shedding of virus-laden droplet and aerosol particles into the air and on the fomite surface and to protect the susceptible worker. Similarly, handwashing and surface disinfection disrupted the indirect tactile transfer of virus to the susceptible worker’s mucous membranes.

The two model outcomes included: 1) the individual and cumulative SARS-CoV-2 infection risks from close contact (droplet and aerosols at 1, 2, 3m), aerosol (<50 µm), and fomite-mediated (droplet and aerosol fallout) exposures to a susceptible worker following an up to 8h-shift with an infected worker in an indoor food manufacturing facility; and 2) the relative reduction in SARS-CoV-2 infection risk attributed to commonly-used infection control interventions (physical distancing, mask use, air change rates, surface disinfection, hand hygiene). The model was developed in R (version 4.0.3; R Development Core Team; Vienna, Austria) using the mc2d package for Monte Carlo simulations and sensitivity analyses (107). For each simulation, 10,000 iterations were run using model parameters selected from defined probability distributions or assigned values (*SI Appendix*, Table S1). Sensitivity analyses conducted on the simulation number demonstrated the risk estimates stabilized after 1,000 iterations, ensuring 10,000 iterations were more than sufficient. Please refer to *SI Appendix* for additional details on the transmission pathways by event.

### 2.3 Data sources

Model parameters derived from the peer-reviewed literature were grouped into five categories and are summarized in *SI Appendix*, Table S1. These included: (i) facility specifications; (ii) viral shedding through cough or breathing events; (iii) fomite-mediated transmission parameters; (iv) dose-response parameters for SARS-CoV-2 infection risk; and (v) risk mitigation interventions (physical distancing, ventilation, mask use, surface disinfection, hand hygiene). The indoor food manufacturing facility simulated in this model was a single-story building (10m x 10m x 10m), representing a total volume of 1000 cubic meters with an internal temperature of 70°F and relative humidity of 40-65%, which is similar to conditions in many facilities in the industry (personal communication with Dr. Sanjay Gummalla, American Frozen Food Industry).

### 2.4 Model validation and calibration

Following model development, validation was conducted to ensure the model structure and risk estimates aligned with the published literature, our evolving understanding of SARS-CoV-2 biology and environmental behavior, and that risk estimates were of an appropriate order of magnitude relative to documented SARS-CoV-2 outbreaks. This involved elicitations of opinions from food processing experts related to the structure of the model and relevant model inputs (e.g. facility ventilation, standard operating procedures for masks, types of surface disinfection products and their frequency of use, etc.); additional details are in *SI Appendix*. Through a systematic review of all published (on or prior to December 2020) SARS-CoV-2 outbreaks with confirmed-positive index and secondary cases, we compared model risk estimates to those derived from real life situations such as SARS-CoV-2 household transmission and recreational or work-based activities with well-defined exposure times and distances. We calibrated our model to the risk for a 1h, close contact (1m) exposure to that of approximately 38% (108, 109), which represents the attack rate from two well-characterized, 1h close contact (1m) exposures. Specifically, this calibration involved adjusting select parameters (e.g. salivary viral titer, coughing frequency, dose-response infectivity), while remaining within the range of parameter values derived from the literature. This calibration was corroborated by additional elicitation of opinions from infectious disease clinicians treating COVID-19 patients in metro-Atlanta.

### 2.5 Aerosol transmission modeling

For aerosol transmission, we leveraged the SARS-CoV-2 air transport model proposed by Zhang *et al*. (79), with modifications as described below and in *SI Appendix*. Briefly, aerosolized particles were assumed to be homogenously distributed throughout the facility, such that aerosol exposure by the susceptible worker was uniform throughout the entire indoor facility. Viral particles were removed through viral decay, based on a temperature- and humidity-specific viral decay rate, λ_v_ (1/s), and removal by ventilation, Q (m^3^/s) converted from air changes per hour (ACH). The baseline model assumed negligible ventilation (ACH 0.1). The total loss of virus-containing room air volume was calculated as:

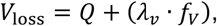

where f_V_ represented the facility volume (m^3^).

The concentration of infectious SARS-CoV-2 particles (PFU/m^3^) at time t remaining in the air, (C_t_), was calculated by:

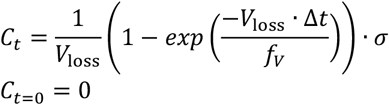

where, σ was the viral shedding rate (PFU/s) from an infected worker and Δ*t* the time change (1/s) from the prior one-hour time step. The virus-laden particles that remained in the air from the prior time-step (based on change in time, Δ*t*), C_t,carry_, (PFU/m^3^) were included in the calculation of the total viral concentration, *C*_*t,total*_, (PFU/m^3^) at time t:

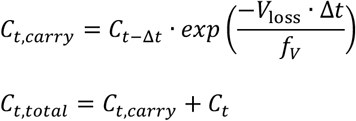

The amount of virus that fell from the air, *Fall*_t,a_ (PFU), at each time-step was determined by the particle size-dependent terminal settling velocity, (m/s) (*𝓋*_*ts*_), the facility surface area (m^2^)(*f*_*a*_), and Δ*t* as follows:

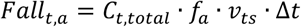

Viral loss due to the aerosol terminal settling velocities and viral decay at 1h intervals was nominal, resulting in accumulation of virus-laden aerosol particles in the air over time.

### 2.6 Close contact transmission modeling

Close contact transmission represented exposure of the susceptible worker within 1, 2, and 3m of the infected worker to droplets (direct spray, respiration) and aerosols (respiration) that deposited into the nasal-pharyngeal region or entered the upper airways. As larger particles (>50 μm) settle out of the air at faster rates than aerosols (<50 μm), for close contact transmission we assumed that all droplets (>50-750 μm) would fall from the air over the course of each one-hour time-step. Therefore, there was no carry-over or accumulation of virus in the air associated with these large droplets between time-steps. To reflect these droplet particle dynamics, particle probability estimates (denoted *pp*) derived from modeling work by Wei *et al*., (110) were used to generate the proportion of droplets that are capable of reaching 1-3m distances as follows:

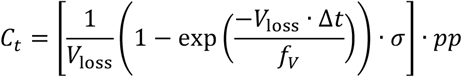

Here, *pp* represented the probability that a droplet would reach 1-3m. This droplet-mediated viral exposure was added to the aerosol viral exposure calculated in section 2.5 to generate the close contact transmission exposures (droplets and aerosols) for 1-3m distances. The inverse of *pp, ppfall*, was used to calculate the viral concentration (PFU/m^3^) fallout from the air and available to contaminate the fomite surface (Fall_t_):

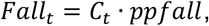

where, *C*_*t*_ is the viral concentration (PFU/m^3^) in the air at time t.

### 2.7 Fomite-mediated transmission modeling

Fomite-mediated transmission was calculated using the droplet and aerosol fallout, Fall_t,a_ (PFU) and Fall_t_ (PFU/m^3^) and the resulting viral contamination on the fomite surface, F_t,a_ and F_t,ccd_ (PFU), at time t:

Fallout from aerosols:

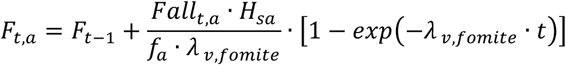

Fallout from close contact droplets:

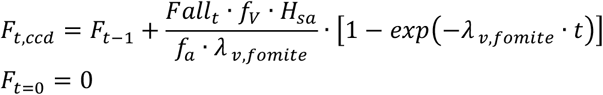

where f_V_ was the volume of the facility (m^3^), H_sa_ was the surface area of the susceptible worker’s hand that touched the fomite surface (m^2^), f_a_ was the cross-sectional area of the facility (m^2^), and λ_v,fomite_ was the viral decay of SARS-CoV-2 on the fomite surface. The concentration of SARS-CoV-2 transferred to a hand, C_hand_ (PFU/h), following a tactile event at time t was calculated using an approach previously applied to influenza A virus exposure (103):

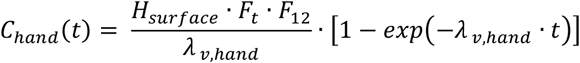

where H_surface_ was the frequency of contacts between the hand and fomite per minute (contacts/min), F_t_ was the viral concentration on the fomite (PFU) at time t, F_12_ was the proportion of virus transferred from fomite to hand, and λ_v,hand_ was the viral decay of SARS-CoV-2 on the hand. The concentration of SARS-CoV-2 on the hand and fallout from close contact droplets both assume first-order loss of virus infectivity.

### 2.8 Risk assessment

Individual and cumulative SARS-CoV-2 infection risks for all three transmission pathways (droplet, aerosol, and fomite-mediated) were estimated for the susceptible worker following an 8h-shift based on exposure to each pathway.

Aerosol and droplet doses were calculated based on the infectious virus concentration in the air (C_t_), lung deposition fraction (L_dep_), inhalation rate (I_R_), and exposure duration (E_t_):

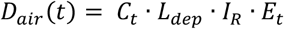

The fomite-mediated dose was calculated from the viral contamination on the hand (C_hand_) at time t, the frequency of hand-to-face contacts (H_face_), the surface area ratio of fingers (F_sa_) to hand (H_sa_), the fraction of pathogens transferred from hand-to-face (F_23_), and the exposure duration (h):

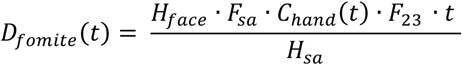

The probability of infection for a given dose based on these individual and combined transmission pathway exposures was estimated using an exponential dose-response model (k_risk_). This model was based on the pooled data from studies of SARS-CoV and murine hepatitis virus infection in mice by intranasal administration (105, 106):

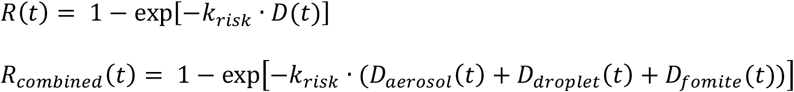

For the combined risk estimate, doses from the individual transmission pathways were assumed to be independent.

### 2.9 Stochastic sensitivity analysis

Spearman rank correlation coefficients were used to identify the most influential parameters in the model by ranking the correlation between risk estimates (8h) and parameter values using the tornado diagram plotting function within the mc2d package in R. To examine the contribution of each parameter to the propagation and overall variability within the model, we applied the built-in “mcratio” function (111) in the mc2d R-package.

### 2.10 Risk mitigation intervention testing

Risk mitigation interventions were selected based on international (EU-OSHA)- and domestic (OSHA, FDA)-recommended guidance and industry practices (according to survey results from food manufacturing facility managers) for worker safety (40) and COVID-19 prevention (37). Interventions included physical distancing (1-3m), concordant universal mask use (surgical, cloth, double masking, N95), improved air changes per hour (ACH 2-8), surface disinfection (3-4 log_10_ virus removal) (112), and hand hygiene (handwashing [2 log_10_ virus removal (113)], alcohol-based hand sanitizer [3 log_10_ virus removal], and glove use [100% virus removal]). Due to the variability in virus removal efficacies of surface disinfectants across products and in hand hygiene practices, we set these efficacies as point values in the model. Surface disinfection was simulated once, twice, four times and hourly per 8h-shift, and hand hygiene was simulated hourly (i.e. handwashing, hand sanitizer, glove use) (personal communications with Dr. Sanjay Gummalla, American Frozen Food Industry). All risk mitigation strategies were assumed to be implemented with 100% compliance and in the manner specified. Parameters associated with the interventions (see Supplementary Materials) were based on controlled laboratory studies.

## Data Availability

Data available upon request.

## Acknowledgments

This work was partially supported by the National Institutes of Health T32 grant (J.S.S., grant 2T32ES012870-16), the National Institute of Food and Agriculture at the U.S. Department of Agriculture (J.S.L. 2018-07410; J.S.S., grant 2020-67034-31728), the National Institute General Medical Sciences (B.A.L R01 GM124280), the National Science Foundation (B.A.L. 2032084), the National Institute Of Allergy And Infectious Diseases of the National Institutes of Health (E.T.S., T32AI138952), and Emory University and the Infectious Disease Across Scales Training Program (IDASTP) (E.T.S). The contents of this paper are solely the responsibility of the authors and do not necessarily represent the official views of the National Institutes of Health, or the U.S. Department of Agriculture.

The authors would like to thank Dr. Sanjay Gummalla (American Frozen Food Industry), Dr. Lory Reveil (American Frozen Food Industry), and Dr. Max Teplitski (Produce Marketing Association) for their valuable time and input as food production and processing experts and for conducting surveys of facilities. We also thank Carol Liu and the infectious disease physicians for their participation in our model calibration efforts.

An open access license has been selected

