## Supplemental Material for "Controlling risk of SARS-CoV-2 infection in essential workers of enclosed food manufacturing facilities"

**This PDF file includes:**

Supplementary text

Figures S1 to S3

Tables S1 to S4

SI References

**Other supplementary materials for this manuscript include the following:**

Not Applicable.

**Supplementary Information Appendix**

**Materials and methods:**

**Model conceptualization and vetting**

Following initial development, the model was vetted by food industry experts: Dr. Sanjay Gummalla [Senior Vice President, Scientific and Regulatory Affairs, American Frozen Food Industry], Dr. Lory Reveil [Director, Scientific and Regulatory Affairs, American Frozen Food Industry], and Dr. Max Teplitski [Chief Science Officer, Produce Marketing Association].

**Transmission pathways by event**

As described, the model was composed of three transmission pathways and we modeled two events, breathing (nasal) to represent asymptomatic or pre-symptom infection with SARS-CoV-2 and cough events (symptomatic individual). The distribution of particle sizes modeled in each transmission pathway were as follows: aerosol transmission of particles with diameters ranging from 0.6-2.2μm (breathing) or 2-50μm (coughing); close-contact transmission ranging from 0.6-

2.2μm (breathing) or 2.0-750μm (coughing) in diameter; and fomite-mediated transmission via contact with a fomite surface, which is contaminated with viral particles based on particle deposition probabilities summed across both aerosol and close contact pathways.

**Data sources**

The model assumed a single infected worker; that the infected and susceptible workers were independent of one another; and that each transmission pathway was independent. For the baseline model, there was negligible ventilation in the facility (defined as air changes per hour (ACH); [ACH 0.1]), however, for the intervention scenarios, ACH values of 2 to 8 were simulated. These corresponded to the number of fresh air exchanges, per hour, resulting in replacement of the entire room air volume. To calculate viral shedding of the infected worker, we converted PCR-based genome equivalent copies to PFU using a 1:100 conversion, as previously applied by Pitol *et al.,* (1). This resulted in SARS-CoV-2 titers in saliva (range: 6.1 to 7.4 $\log_{10}$ PFU) (2, 3), representative of peak virus titer reported within the first week of symptom onset (3, 4) and the acute phase of infection, when the majority of transmission events are thought to occur (4). The same distribution of shedding data was used for symptomatic and asymptomatic individuals as these are not known to be statistically different (5). To determine the amount of virus expelled into the air by the infected worker, the total fraction of saliva volume released during coughing or breathing was calculated for each droplet (50-60μm, 60-100μm, >100μm) and aerosol (<50μm) range as described in (6) using respiratory particle counts and size distributions from empirical studies (7). Viral decay (8) was included at three stages in the model: 1) virus-containing aerosols or droplets in the air; 2) virus-contamination on a fomite surface; and 3) the virus-contaminated hands of the susceptible worker. For fomite-mediated transmission, virus transfer efficiencies from fomite surfaces to hands leveraged laboratory-based studies using the viral surrogate MS2 (9). Sequential tactile events were modeled from the initial contact of the susceptible worker's hand to the fomite surface (one contact/minute) followed by hand contact to facial mucous membranes (0.8 contacts/minute). SARS-CoV-2 infection risks were estimated using an exponential dose-response model based on the pooled data from studies of SARS-CoV and murine hepatitis virus infection in mice by intranasal administration (10, 11) with the $ID_{50}$ equal to 102 infectious particles

Laboratory-based studies on mask filtration efficacies (12-15) were used to evaluate the impact of cloth, surgical, double masking (surgical followed by cloth), and N95 respirators. Using these empirical studies, which reported mask filtration efficacies for either source control and/or recipient protection, we calculated the mask efficiencies for when the infected worker wore the mask (source) and for when the susceptible worker (recipient) wore the mask. Hand hygiene and surface disinfection virus removal efficacies were representative of current CDC and EPA List N: Disinfectants for Coronavirus (COVID-19) products: hand washing (2 $\log_{10}$ virus removal) (16), alcohol-based hand sanitizer (3 $\log_{10}$ virus removal), surface disinfection (3-4 $\log_{10}$ virus removal) (17). For all mitigation strategies (mask use, hand hygiene, and surface disinfection), we assumed that these were implemented with 100% compliance and in the specified manner.

**Air transport modeling:**

To simulate virus-laden aerosols released from the infected worker through either breathing or coughing, the total viral shedding was calculated as:

$$E_{virus} = V_F \cdot Freq_E \cdot 10^{(\log_{10} C_{virus})}$$

Here, $E_{virus}$ represents the total virus shed per hour (PFU/h), $V_F$ the fraction of volume associated with the particle sizes (μm) per respiratory event, $Freq_E$ the respiratory event frequency per hour, and $C_{virus}$ the infectious virus concentration in saliva (PFU/mL). $E_{virus}$ was then converted to σ, viral shedding rate (PFU/s).

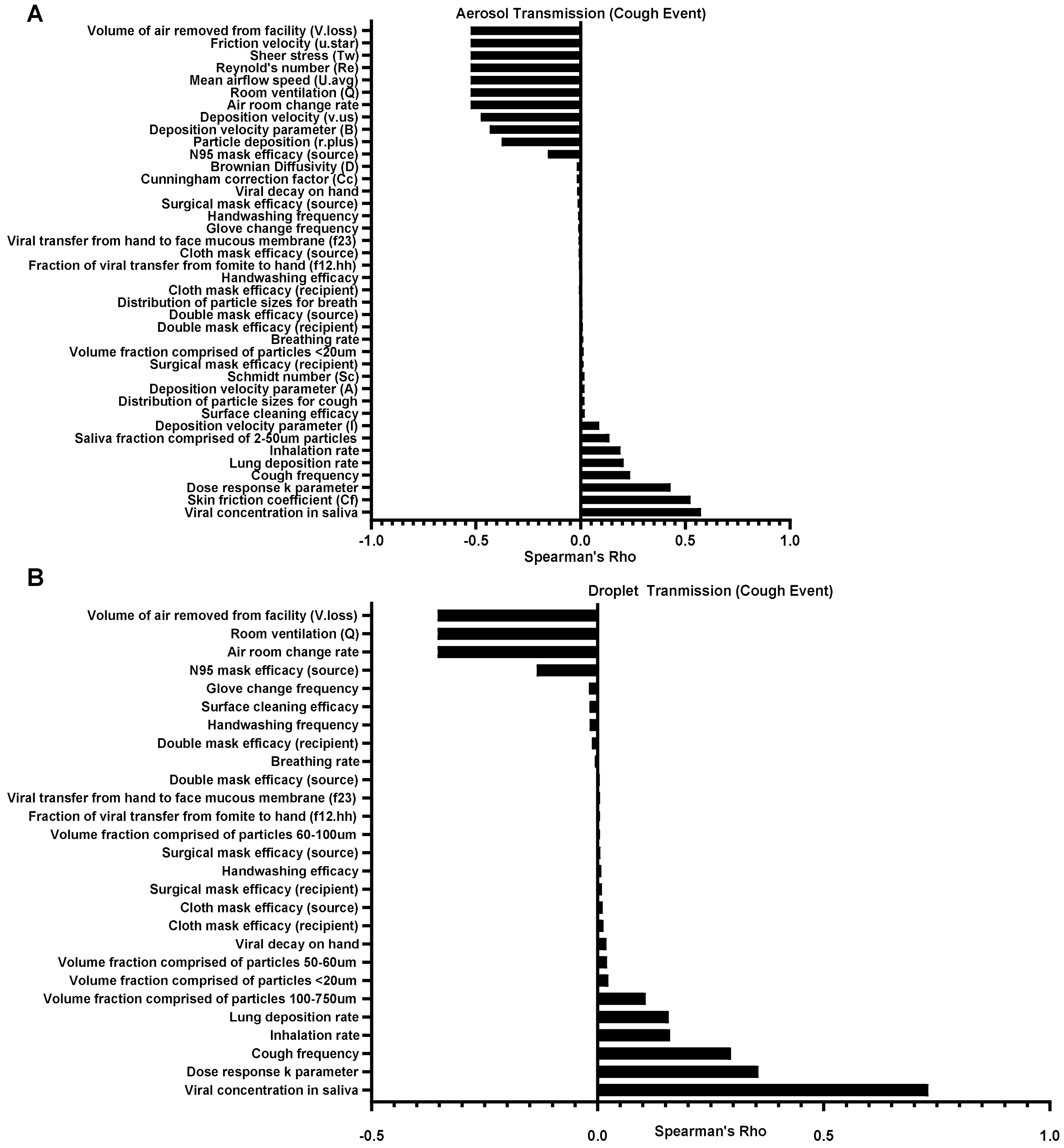


**Fig. S1.** Tornado plots for SARS-CoV-2 cumulative infection risk following an 8h exposure with all interventions included. Spearman rank correlation coefficients were used to assess the most influential parameters in the model. To represent the mask intervention, N95 ventilators were selected as these had the highest filtration efficiency.

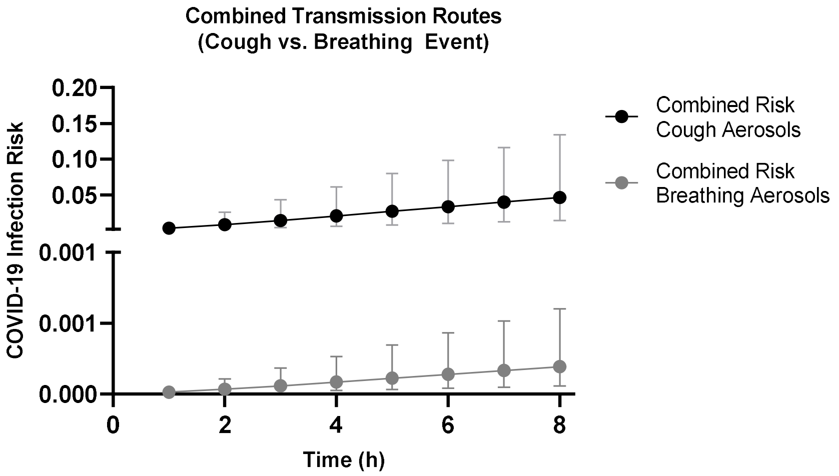


**Fig. S2.** Comparison of combined infection risks resulting from exposures (1 to 8h) to aerosols (<50 μm) originating from a symptomatic coughing infected worker relative to an asymptomatic breathing infected worker in the absence of any interventions.

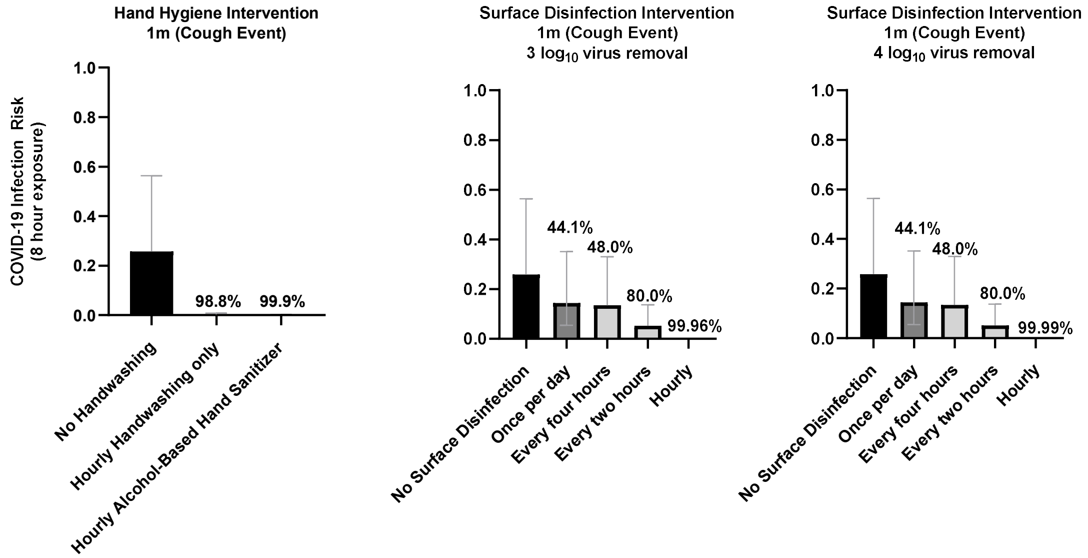


**Fig. S3.** Impact of hand hygiene (2 $log_{10}$ virus removal handwashing efficacy; 3 $log_{10}$ virus removal alcohol-based hand sanitizer efficacy) and surface disinfection (3 $log_{10}$ or 4 $log_{10}$ virus removal efficacy) interventions on isolated fomite-mediated infection risk following 8h cumulative exposure to an infected worker (coughing) at a 1m distance. The included percentages above each column represent the percent reduction relative to no intervention (e.g. no handwashing or no surface disinfection).

**Table S1.** Model parameter inputs and distributions.

| Parameter | Units | Description | Distribution | Input Values | Citations |
|---|---|---|---|---|---|
| **Facility specifications** | | | | | |
| $F_{Dimensions}$ | m | Dimensions of the facility | Point value | 10m x 10m x 10m | Assumed |
| $F_{volume}$ | $m^3$ | Volume of the facility | Point value | 1000 cubic meters | Calculated |
| $f_a$ | $m^2$ | Surface area of facility | Point value | 100 | Calculated |
| $F_{Temperature}$ | Degrees Fahrenheit | Facility ambient temperature | Point value | 70 | Assumed |
| $k_v$ | $m^2$/second | Kinematic viscosity of facility air for 70°F | Point value | 1.52E-5 | (18) |
| $d_v$ | N*s/$m^2$ | Dynamic viscosity of air at 70°F | Point value | 1.82E-5 | (18) |
| $\rho_{air}$ | kg/$m^3$ | Air density at sea level | Point value | 1.225 | (18) |
| k | J/K | Boltzmann's constant | Point value | 1.38E-23 | (18) |
| $\lambda_{air}$ | cm | Mean free path of air at sea level | Point value | 3.4E-6 | (18) |
| **Viral shedding** | | | | | |
| $Log_{10}(C_{virus})$ | PFU/mL | Concentration of virus in saliva | Triangular | 6.8 (6.1, 7.4) | (2, 3) |
| $d_{p,c}$ | cm | Diameter of respiratory particles for coughing event | Triangular | 6E-4 (2E-4, 4.9E-3) | (7) |
| $d_{p,b}$ | cm | Diameter of respiratory particles for breathing event | Triangular | 8E-5 (3E-5, 2E-3) | (19, 20) |
| $V_{F,c}$ | mL/Cough | Fraction of volume associated with aerosols (2–45µm) | Triangular | 2.3E-6 (1.4E-6, 2.6E-6) | (7) |
| $V_{F,c}$ | mL/Cough | Fraction of volume associated with droplets (50–60µm) | Triangular | 6.0E-6 (3.5E-6, 6.7E-6) | (7) |
| $V_{F,c}$ | mL/Cough | Fraction of volume associated with droplets (60–100µm) | Triangular | 4.9E-6 (1.1E-6, 8.4E-6) | (7) |
| $V_{F,c}$ | mL/Cough | Fraction of volume associated with droplets (100–750µm) | Triangular | 6.8E-3 (4.0E-3, 7.6E-3) | (7) |
| $V_{F,b}$ | mL/Breath | Fraction of volume associated with aerosols (0.6-2.2µm) | Uniform | (1.1E-10, 2.9E-10) | (19, 20) |
| $F_C$ | Cough/h | Number of coughs per hour | Triangular | 25 (10, 39.25) | (21, 22) |
| $F_B$ | Breaths/h | Number of breaths per hour | Uniform | (960, 1,200) | (23) |
| $\lambda_{virus}$ | Hour | Viral decay of SARS-CoV-2 at 40% relative humidity, 70°F | Point value | 0.614 | (8) |

| | | | | | |
|---|---|---|---|---|---|
| pp | Probability | Probability droplets will reach a specified distance (m) | Point value | 50-60μm:<br>1m: 0.82, 2m: 0.43, 3m: 0.19<br>60-100μm:<br>1m: 0.44, 2m: 0.01;<br>>100μm:<br>1m: 0.04 | (24) |
| **Risk mitigation interventions[1]** | | | | | |
| $S_{mask}$ | Log reduction | Source protection surgical mask efficacy | Uniform | (0.39, 0.57) | (14, 25, 26) |
| $C_{mask}$ | Log reduction | Source protection cloth mask efficacy | Uniform | (0.31, 0.62) | (14, 25, 26) |
| $N95_{mask}$ | Log reduction | Source protection N95 mask efficacy | Uniform | (1.89, 2.15) | (14, 25, 26) |
| $D_{mask}$ | Log reduction | Double masking (surgical followed by cloth) efficacy | Uniform | (0.27, 1.89) | (15) |
| $RS_{mask}$ | Percent reduction | Recipient surgical mask efficacy | Uniform | (0.37, 0.998) | (14, 25, 26) |
| $RC_{mask}$ | Percent reduction | Recipient cloth mask efficacy | Uniform | (0.17, 0.887) | (14, 25, 26) |
| $RN95_{mask}$ | Percent reduction | Recipient N95 mask efficacy | Point value | 0.79 | (14, 25, 26) |
| $RD_{mask}$ | Percent reduction | Recipient double masking (surgical followed by cloth) efficacy | Uniform | (0.40, 0.9685) | 42] |
| $SC_{eff}$ | Log reduction | Surface disinfection efficacy | Point value | $3Log_{10}$ virus | (17) |
| $HW_{eff}$ | Log reduction | Hand washing efficacy | Point value | $2Log_{10}$ virus | (27, 28) |
| $HW_{freq}$ | Handwashing /h | Frequency of handwashing per hour | Point value | 1.0 | Expert elicitation |
| $G_{freq}$ | Glove changes/h | Frequency of glove changes per hour | Point value | 1.0 | Expert elicitation |
| $F_{24}$ | Percent reduction | Viral removal efficacy attributed to glove changes | Point value | 1.0 | (29) |
| $R_{air}$ | Air changes/h | Frequency of room air changes per hour (ACH) | Point value | Base model: 0.1<br>Intervention: 2-8 | Expert elicitation |
| **Fomite-mediated transmission** | | | | | |
| $F_{sa}$ | $m^2$ | Fomite surface area representative of 0.5m x 0.5m | Point value | 0.25 | Assumed |
| $Fingers_{sa}$ | $m^2$ | Surface area of three finger tips touching the surface | Point value | 0.00042 | (30) |
| $H_{sa}$ | $m^2$ | Area of two hands (palms only) | Point value | 0.049 | (30) |
| $F_{decay}$ | Hour | Viral decay rate (PFU per hour) | Point value | 0.15 | (8) |
| $F_{12}$ | PFU | Viral transfer fraction from fomite to hand | Normal | 0.374 (0.16) | (31) |

| | | | | | |
|---|---|---|---|---|---|
| | | with relative humidity (40-65%) | | | |
| $F_{21}$ | PFU | Viral transfer fraction from hand to fomite surface | Point value | 0.025 | (32-35) |
| $F_{23}$ | PFU | Viral transfer fraction from hand to face | Normal | 0.20 (0.063) | (31) |
| freq.hs | Contacts/min | Frequency of contacts from hand to fomite | Point value | 1 | Assumed |
| freq.hf | Contacts/min | Frequency of contacts from hand to face | Point value | 0.8 | (36) |
| $Hand_{decay}$ | Minutes | Viral decay rate on hands (PFU/min) | Uniform | (0.92, 1.47) | (33) |
| **SARS-CoV-2 dose and risk characterization** | | | | | |
| $L_{dep}$ | PFU | Deposition fraction of infectious virus into the lungs (Upper and lower respiratory tract; facial mucous membranes) | Point value | 1 | Assumed; (37) |
| $I_R$ | $m^3$ / h | Inhalation rate per hour representing moderate activity | Uniform | (1.62, 3.18) | (38) |
| $k_{risk}$ | No units | Dose-response parameter | Point value | 0.00680 | (1) |

[1]All interventions were assumed to be implemented with 100% compliance.

**Table S2.** Relative reduction (as a percentage) in combined risk from close contact transmission (droplet and aerosol) as a function of distance from an infected worker (coughing) and exposure time.

| Time (h) | 3m versus 2m | 2m versus 1m | 3m versus 1m |
|---|---|---|---|
| **1** | 35.0% | 97.3% | 98.4% |
| **2** | 35.0% | 96.2% | 97.5% |
| **3** | 36.1% | 94.9% | 96.7% |
| **4** | 37.7% | 93.3% | 95.8% |
| **5** | 39.3% | 91.5% | 94.8% |
| **6** | 40.8% | 89.4% | 93.7% |
| **7** | 42.0% | 87.0% | 92.5% |
| **8** | 43.2% | 84.3% | 91.1% |

**Table S3.** The contribution of individual parameter variability to the overall model variability as assessed by the ratio of the 97.5$^{th}$ model estimate/the median model estimate stratified by transmission pathway (aerosol, droplet, and fomite-mediated).

| Parameter | Variability Ratio[1] Aerosol and Aerosol-Mediated Fomite Pathways | Variability Ratio[1] Droplet and Droplet-Mediated Fomite Pathways |
|---|---|---|
| Virus concentration in saliva (PFU/mL) | 1.1 | 1.1 |
| Distribution of particle sizes for cough (μm) | 2.5 | N/A[2] |
| Distribution of particle sizes for breath (μm) | 2.7 | N/A |
| Saliva fraction comprised of 2-50um particles (mL/cough event) | 1.2 | N/A |
| Volume fraction comprised of particles <20um generated from breathing (mL/breathing event) | 1.4 | 1.4 |
| Volume fraction comprised of particles 50-60um (mL/cough event) | N/A | 1.2 |
| Volume fraction comprised of particles 60-100um (mL/cough event) | N/A | 1.6 |
| Volume fraction comprised of particles 100-750um (mL/cough event) | N/A | 1.2 |
| Breathing rate (breaths/h) | 1.1 | 1.1 |
| Cough frequency (coughs/h) | 1.4 | 1.6 |
| Surgical mask efficacy (source) (log virus reduction) | 1.2 | 1.2 |
| Cloth mask efficacy (source) (log virus reduction) | 1.3 | 1.3 |
| N95 mask efficacy (source) (log virus reduction) | 1.1 | 1.1 |
| Double mask efficacy (source) (log virus reduction) | 1.7 | 1.7 |
| Surgical mask efficacy (recipient) (percent reduction) | 1.4 | 1.4 |
| Cloth mask efficacy (recipient) (percent reduction) | 1.6 | 1.6 |
| Double mask efficacy (recipient) (percent reduction) | 1.4 | 1.4 |
| Virus shedding concentration (PFU/h) | 3.6 | 5.5 |
| Air room change rate ($m^3$/h) | 1.9 | 1.9 |
| Room ventilation (Q) ($m^3$/s) | 1.9 | 1.9 |
| Mean airflow speed (U.avg) (m/s) | 1.9 | N/A |
| Reynold's number (Re) (dimensionless) | 1.9 | N/A |
| Skin friction coefficient (Cf) (dimensionless) | 1.4 | N/A |
| Sheer stress (Tw) (N/$m^2$) | 3.3 | N/A |
| Friction velocity (u.star) (m/s) | 1.8 | N/A |
| Particle deposition (r.plus) (dimensionless) | 4.2 | N/A |
| Cunningham correction factor (Cc) (dimensionless) | 1.0 | N/A |
| Brownian Diffusivity (D) ($m^2$/s) | 4.2 | N/A |
| Schmidt number (Sc) (dimensionless) | 2.5 | N/A |
| Deposition velocity parameter (A) (dimensionless) | 1.0 | N/A |
| Deposition velocity parameter (B) (dimensionless) | 0.9 | N/A |
| Deposition velocity parameter (I) (dimensionless) | 1.8 | N/A |
| Deposition velocity (v.us) (m/s) | 2.4 | N/A |
| Volume of air removed from facility (V.loss) ($m^3$/s) | 1.7 | 1.8 |
| Virus concentration in the air 1h (PFU/$m^3$) | 5.1 | 7.1 |

| | | |
|---|---|---|
| Virus concentration in the air 2h (PFU/m$^3$) | 5.9 | 7.1 |
| Virus concentration in the air 3h (PFU/m$^3$) | 6.3 | 7.1 |
| Virus concentration in the air 4h (PFU/m$^3$) | 6.4 | 7.1 |
| Virus concentration in the air 5h (PFU/m$^3$) | 6.5 | 7.1 |
| Virus concentration in the air 6h (PFU/m$^3$) | 6.5 | 7.1 |
| Virus concentration in the air 7h (PFU/m$^3$) | 6.5 | 7.1 |
| Virus concentration in the air 8h (PFU/m$^3$) | 6.5 | 7.1 |
| Virus fallout from air (fallout1)[3] (PFU) | 4.0 | 7.7 |
| Virus fallout from air (fallout2)[3] (PFU) | 4.1 | 7.7 |
| Virus fallout from air (fallout3)[3] (PFU) | 4.3 | 7.7 |
| Virus fallout from air (fallout4)[3] (PFU) | 4.3 | 7.7 |
| Virus fallout from air (fallout5)[3] (PFU) | 4.3 | 7.7 |
| Virus fallout from air (fallout6)[3] (PFU) | 4.3 | 7.7 |
| Virus fallout from air (fallout7)[3] (PFU) | 4.3 | 7.7 |
| Virus fallout from air (fallout8)[3] (PFU) | 4.3 | 7.7 |
| Surface cleaning efficacy (log virus reduction) | 1.0 | 1.0 |
| Handwashing efficacy (log virus reduction) | 1.0 | 1.0 |
| Handwashing frequency (Handwashing/h) | 1.7 | 1.7 |
| Glove change frequency (Glove changes/h) | 1.7 | 1.7 |
| Fraction of virus transfer from fomite to hand (f12.hh) (PFU) | 1.8 | 1.8 |
| Virus transfer from hand to face mucous membrane (f23) (PFU) | 1.6 | 1.6 |
| Viral decay on hand (h) | 1.2 | 1.2 |
| Virus concentration on hand 1h-8h (<50um) (PFU) | 0.0 | 0.0 |
| Virus concentration on hand 1h (PFU) | N/A | 86060-86348 |
| Virus concentration on hand 2h (PFU) | N/A | 86771-86171 |
| Virus concentration on hand 3h (PFU) | N/A | 86975-86116 |
| Virus concentration on hand 4h (PFU) | N/A | 86464-86818 |
| Virus concentration on hand 5h (PFU) | N/A | 86239-87562 |
| Virus concentration on hand 6h (PFU) | N/A | 85849-87101 |
| Virus concentration on hand 7h (PFU) | N/A | 85854-87198 |
| Virus concentration on hand 8h (PFU) | N/A | 86021-87302 |
| Dose transferred from hand to face h1 (DT.hh1) (PFU) | 0.0 | 93872 - 96608 |
| Dose transferred from hand to face h2 (DT.hh2) (PFU) | 0.0 | 93582-96453 |
| Dose transferred from hand to face h3 (DT.hh3) (PFU) | 0.0 | 93230 - 95814 |
| Dose transferred from hand to face h4 (DT.hh4) (PFU) | 0.0 | 92680-94779 |
| Dose transferred from hand to face h5 (DT.hh5) (PFU) | 0.0 | 92870-93519 |
| Dose transferred from hand to face h6 (DT.hh6) (PFU) | 0.0 | 93105-93220 |
| Dose transferred from hand to face h7 (DT.hh7) (PFU) | 0.0 | 93030-93065 |
| Dose transferred from hand to face h8 (DT.hh8) (PFU) | 0.0 | 90158-91670 |
| Lung deposition rate (PFU) | 1.3 | 1.3 |
| Inhalation rate (m$^3$/h) | 1.3 | 1.3 |
| Dose h1 (total PFU) | 5.5 | 7.78-8.69 |
| Dose h2 (total PFU) | 6.4 | 7.78-8.69 |

| | | |
|---|---|---|
| Dose h3 (total PFU) | 6.8 | 7.78-8.69 |
| Dose h4 (total PFU) | 6.9 | 7.78-8.69 |
| Dose h5 (total PFU) | 7.0 | 7.78-8.69 |
| Dose h6 (total PFU) | 7.0 | 7.78-8.69 |
| Dose h7 (total PFU) | 7.0 | 7.78-8.69 |
| Dose h8 (total PFU) | 7.0 | 7.78-8.69 |
| Fomite concentration h1 (total PFU) | 4.0 | 7.7 |
| Fomite concentration h2 (total PFU) | 4.1 | 7.7 |
| Fomite concentration h3 (total PFU) | 4.1 | 7.7 |
| Fomite concentration h4[4] (total PFU) | 4.2 | 10.4 |
| Fomite concentration h5 (total PFU) | 4.2 | 7.7 |
| Fomite concentration h6 (total PFU) | 4.3 | 7.7 |
| Fomite concentration h7 (total PFU) | 4.3 | 7.7 |
| Fomite concentration h8[4] (total PFU) | 4.3 | 10.4 |
| Dose-response k parameter (dimensionless) | 1.6 | 1.6 |
| Aerosol/Droplet risk cumulative h1 | 6.8 | 9.15-10.17 |
| Aerosol/Droplet risk cumulative h2 | 7.3 | 9.15-10.17 |
| Aerosol/Droplet risk cumulative h3 | 7.5 | 9.15-10.17 |
| Aerosol/Droplet risk cumulative h4 | 7.8 | 9.15-10.17 |
| Aerosol/Droplet risk cumulative h5 | 7.9 | 9.15-10.17 |
| Aerosol/Droplet risk cumulative h6 | 8.0 | 9.15-10.17 |
| Aerosol/Droplet risk cumulative h7 | 8.0 | 9.15-10.17 |
| Aerosol/Droplet risk cumulative h8 | 8.1 | 9.15-10.17 |
| Fomite cumulative risk h1 | 0.0 | 110857-113244 |
| Fomite cumulative risk h2 | 0.0 | 110276-112724 |
| Fomite cumulative risk h3 | 0.0 | 111086-113010 |
| Fomite cumulative risk h4 | 0.0 | 110960-112044 |
| Fomite cumulative risk h5 | 0.0 | 109548-111880 |
| Fomite cumulative risk h6 | 0.0 | 108923-111936 |
| Fomite cumulative risk h7 | 0.0 | 108258-111603 |
| Fomite cumulative risk h8 | 0.0 | 107974-111504 |

[1]The variability ratio is dimensionless. This ratio represents the 97.5$^{th}$ model estimate/the median model estimate for each parameter to determine the relative contribution to the overall variability in the model. Lower values represent less variability. This analysis used the “mcratio” function in the mc2d package in R.

[2]N/A indicates not applicable. For instance the volume fraction comprised of particles > 50 μm would not be included in the aerosol transmission pathways as we considered particles 50 μm or less to be aerosols.

[3]Virus fallout was defined as virus-laden particles that were removed from the air due to their terminal settling velocity or gravitational forces. For the droplet pathway, particle fallout was composed of droplets 50-100 μm in diameter. Droplets 100-750 μm were not included in the fallout calculation as these were assumed to fall out of radius of the fomite.

[4]Surface disinfection set for 4h and 8h for the sensitivity analysis.

Note: all cumulative risks (aerosol, droplet, fomite-mediated) are dimensionless and represent the probability of infection, bounded between 0 and 1, of a fully susceptible worker following exposure to a single infected worker for 1h to a cumulative 8h duration.

**Table S4.** Population-level $R_0$[1] values derived from SARS-CoV-2 combined transmission risk estimates.

| Time (h) | Combined Risk 1m | | | Combined Risk 2m | | | Combined Risk 3m | | | Combined Risk Aerosol | | |
|---|---|---|---|---|---|---|---|---|---|---|---|---|
| | **Median** | **2.50%** | **95%** | **Median** | **2.50%** | **95%** | **Median** | **2.50%** | **95%** | **Median** | **2.50%** | **95%** |
| 1 | 2.38 | 0.92 | 5.00 | 0.06 | 0.03 | 0.14 | 0.04 | 0.02 | 0.09 | 0.02 | 0.01 | 0.07 |
| 2 | 3.97 | 1.76 | 6.43 | 0.15 | 0.06 | 0.33 | 0.10 | 0.04 | 0.22 | 0.06 | 0.02 | 0.18 |
| 3 | 5.04 | 2.54 | 6.84 | 0.26 | 0.11 | 0.55 | 0.16 | 0.07 | 0.37 | 0.10 | 0.03 | 0.30 |
| 4 | 5.75 | 3.23 | 6.96 | 0.38 | 0.17 | 0.81 | 0.24 | 0.10 | 0.53 | 0.14 | 0.04 | 0.43 |
| 5 | 6.22 | 3.86 | 6.99 | 0.53 | 0.23 | 1.11 | 0.32 | 0.13 | 0.71 | 0.19 | 0.06 | 0.56 |
| 6 | 6.51 | 4.41 | 7.00 | 0.69 | 0.31 | 1.45 | 0.41 | 0.17 | 0.89 | 0.23 | 0.07 | 0.69 |
| 7 | 6.71 | 4.90 | 7.00 | 0.87 | 0.39 | 1.81 | 0.51 | 0.21 | 1.08 | 0.28 | 0.09 | 0.81 |
| 8 | 6.83 | 5.32 | 7.00 | 1.07 | 0.48 | 2.21 | 0.61 | 0.26 | 1.28 | 0.32 | 0.10 | 0.94 |

[1]$R_0$ is defined as the basic reproductive number, which represents the expected number of new cases from one infected individual in a fully susceptible population.
